# Stepwise evolution of carbapenem-resistance, captured in patient samples and evident in global genomics of *Klebsiella pneumoniae*

**DOI:** 10.1101/2021.06.21.21259170

**Authors:** Laura Perlaza-Jiménez, Jonathan J. Wilksch, Christopher J. Stubenrauch, Tao Chen, Yajie Zhao, Tieli Zhou, Trevor Lithgow, Vijaykrishna Dhanasekaran

## Abstract

The World Health Organization ranks *Klebsiella pneumoniae* as a priority antimicrobial-resistant (AMR) pathogen requiring urgent study. New strategies for diagnosis and treatment, particularly for those *Klebsiella* that are classified as carbapenem-resistant Enterobacteriaceae (CRE) need to recognize the increased prevalence of non-carbapenemase producing CRE (non-CP CRE). By integrating diverse *Klebsiella* genomes with known CRE phenotypes, we successfully identified a synchronized presence of CRE phenotype-related genes in plasmids and chromosomes in comparison to strains with carbapenem susceptible phenotypes. The data revealed a major contribution to CRE comes from the combined effect of chromosome and plasmid genes potentiated by modifications of outer membrane porins. Our computational workflow identified key gene contributors to the non-CP CRE phenotype, including those that lead to an increase of antibiotic expulsion by enhanced efflux pump activity and mobile elements that reduce antibiotic intake, such as IS*1* and Tn*3*-like elements. These findings are consistent with a new model wherein a change to the balance in drug influx and efflux potentiates the ability of some beta-lactamases to enable survival in the presence of carbapenems. Analysis of the large numbers of documented CRE infections, as well as forensic analysis of a case study, showed that this potentiation can occur in short timeframes to deliver a non-CP CRE infection. Our results suggest that the multiple genes that function to build an AMR phenotype can be diagnosed, so that strains that will resist treatment with carbapenem treatment will be evident if a comprehensive genome-based diagnostic for CRE considers all of these sequence-accessible features.

**SIGNIFICANCE:** Carbapenem-resistant Enterobacteriaceae (CRE) has emerged as an important challenge in health-care settings, with *Klebsiella pneumoniae* playing a major role in the global burden of CRE infections. Through systematic characterisation of the chromosome and plasmid genes of *K. pneumoniae* strains and their antimicrobial traits we identified new CRE mechanisms that are important for accurate diagnosis of carbapenem-resistant AMR. The development of comprehensive genomics-based diagnostics for CRE will need to consider the multiple gene signatures that impact together to deliver non-carbapenemase, carbapenem-resistant infections.

## Introduction

*Klebsiella pneumoniae* is a priority antimicrobial-resistant (AMR) pathogen requiring urgent study and new strategies for diagnosis and treatment (1). While carbapenems had stood as a last-line treatment for *Klebsiella* infections, the rise in carbapenem-resistant clones of *Klebsiella* species, particularly prevalent across Asia, pose a rapidly growing threat. *K. pneumoniae* gain carbapenem resistance by expression of carbapenemases that hydrolyse carbapenems and other beta-lactam antibiotics. Acquisition of plasmid-encoded carbapenemases converts carbapenem sensitive strains of *K. pneumoniae* to resistant strains with what appears to be alarming ease (2, 3). Recently, a more complicated non-carbapenemase (non-CP) carbapenem-resistant phenotype has been recognized globally, where a prospective, multicentre, cohort study for carbapenem-resistant Enterobacteriaceae (CRE) in USA revealed that 41% of the isolates did not encode carbapenemases (4). To improve options to control AMR, a better understanding of the non-CP mechanisms and their relative incidence is imperative.

Beta-lactam antibiotics such as carbapenems target the bacterial periplasm. These drugs enter the periplasm via porins in the bacterial outer membrane, and can be secreted by efflux pumps along the outer membrane (5). As a result, a key non-CP CRE mechanism in strains that express an extended spectrum beta-lactamase (ESBL), is either loss-of-function mutations in outer membrane porins to diminish beta-lactam entry (6, 7), and/or overproduction of efflux pumps to increase drug export from the periplasm (8, 9). Together, genotypes such as this result in a decreased concentration of carbapenem in the periplasm, which if sufficiently low, can be cleared by the action of ESBLs to generate a non-CP CRE phenotype (6). Consistent with this, we recently reported a case study where a CRE phenotype was ultimately generated by loss of porin function in a strain that expressed no recognizable carbapenemase, but which carried the *bla*_DHA-1_ gene encoding an ESBL (10).

Evidence from bacterial population biology suggests that a positive epistasis between the chromosomal genes and plasmids enables long-term survival (11). In many cases variability in resistance to antimicrobials is the result the action of a combination of genes with different, independent functions (12); however, these associations between chromosomal and plasmid genes to enable AMR phenotypes remains understudied. There is an emerging acceptance for whole-genome sequencing (WGS)-based strategies to predict AMR phenotypes down to the detail of minimal inhibitory concentrations (MICs) for specific drugs (13), as they reduce the time needed for diagnosis to inform therapy and improve patient outcomes. Diagnosis from rapidly acquired genome sequence data is a promising new approach, where WGS data can be used as a basis for machine learning methods to deliver insight into genomic features that are involved in AMR (14) and this approach has been applied in at least one study to predict MIC data for clinical isolates of *Klebsiella* (15).

Here we identified genes that act in association to generate CRE phenotypes, by systematically analysing *Klebsiella pneumoniae* strains with clinical and phenotypic characterisation and genomic data with accurate chromosome and plasmid gene mapping. In our analysis we observed three plasmid-mediated mechanisms that contribute to carbapenem resistance: (i) strains with plasmids that carry carbapenemase producing genes, (ii) strains with plasmids that carry beta-lactamase genes and chromosomes with defective porins and (iii) modifications in the membrane functions that increase the efflux of antibiotics, either by changes in the regulation of efflux pump expression or the duplication of efflux pumps genes. During this analysis, we discovered drug-resistance in non-CP strains could be affected through genes encoding efflux pumps. When analyzing gene associations between plasmids and chromosomal genes, we found an enrichment of membrane component genes, suggesting that a complementary balance between influx and efflux is necessary for resistance. The presence of mobile elements was associated with the resistant phenotype, suggesting an evolutionary trend and non-random mobility of plasmids.

## RESULTS

### Within-host evolution of a CRE infection caused by *K. pneumoniae*

Three *K. pneumoniae* isolates (FK-2624, FK-2723 and FK-2820) collected from sputum samples of a hospitalized patient across a period of 113 days (10) showed phenotypic evidence of within-host evolution of carbapenem resistance (Fig. 1a). To test their suggested relatedness, the genomes of these three isolates were sequenced. Phylogenies generated comparing these strains with all publicly available completed *K. pneumoniae* genomes showed that the three in-host isolates were most closely related to each other (Fig. 1b). A sequence identity of >98% was observed for FK-2624, FK-2723 and FK-2820, but with only 307 single nucleotide variants (SNVs) (∼ 0.01% core genome variability) using FK-2624 as reference. The remainder of the 2% variation between genomes was attributed to the increased lengths of the assembled genomes (from 5700 kb for FK-2624 on day 26, to 6053 kb for FK-2723 on day 71, to 6057 kb for FK-2820 on day 113) (Supplementary Table 1). The estimated SNVs between the genomes was significantly less than the expected ∼2628 SNVs based on a mutation rate of 1 × 10^−7^ per nucleotide site per generation. Further, when considering the third isolate FK-2820 as reference, 82 SNVs were shared between the first two isolates, FK-2624 and FK-2723. Taken together, these results strongly support the within-host emergence of FK-2820 from FK-2723 which likewise arose from the progenitor FK-2624 upon antibiotic use (Fig. 1a).

**Figure 1.**
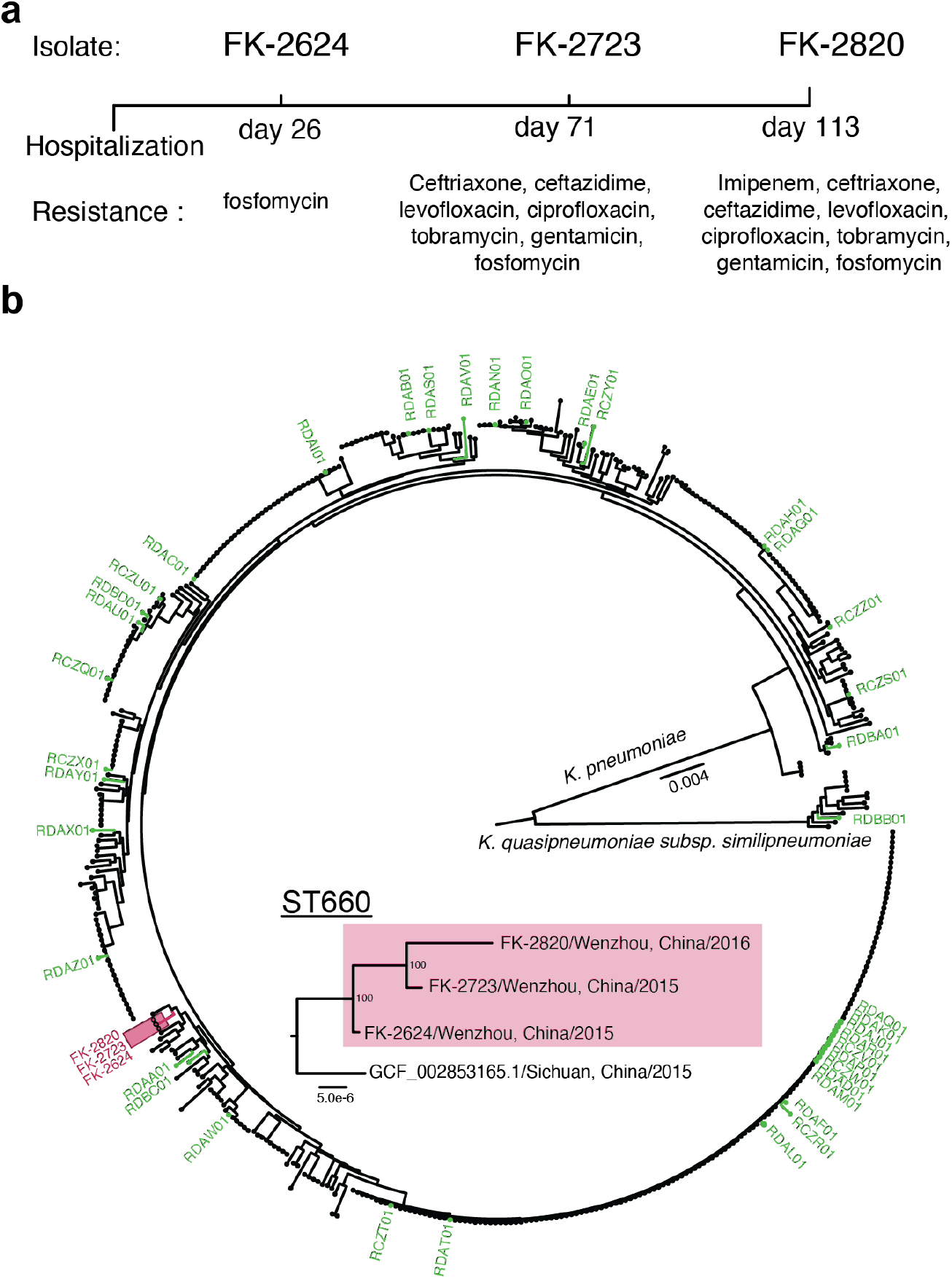
A case-study of within-host emergence of non-CP CRE. **a**, Timeline of K. pneumoniae isolate collection and their antimicrobial properties. For detailed patient clinical and treatment history, see ref. 15. **b**, Phylogenetic relationships linking FK-2624, FK-2723 and FK-2820 with 597 complete genomes of K. pneumoniae and K. quasipneumoniae. Scale bars represents nucleotide substitutions per site. Red square highlights the Wenzhou strains. Strains subjected to further study are shown in green.

To understand the nature of the changes in FK-2820 that yield the CRE phenotype (Fig. 2a), physical maps of the genomes were inferred from the WGS data. Genomic analysis using a plasmid predictor (mlplasmids) (16) showed that the three isolates contained two putative plasmids. A smaller plasmid of 211 kb was found in all three isolates. Due to SNVs and gene content, the plasmids have unique names: pTC1-2624, pTC1-2723 and pTC1-2820. A larger plasmid of 340 kb was found in the latter two strains FK-2723 and FK-2820, denoted as pTC2-2723 and pTC2-2820, respectively (Fig. 2b).

**Figure 2.**
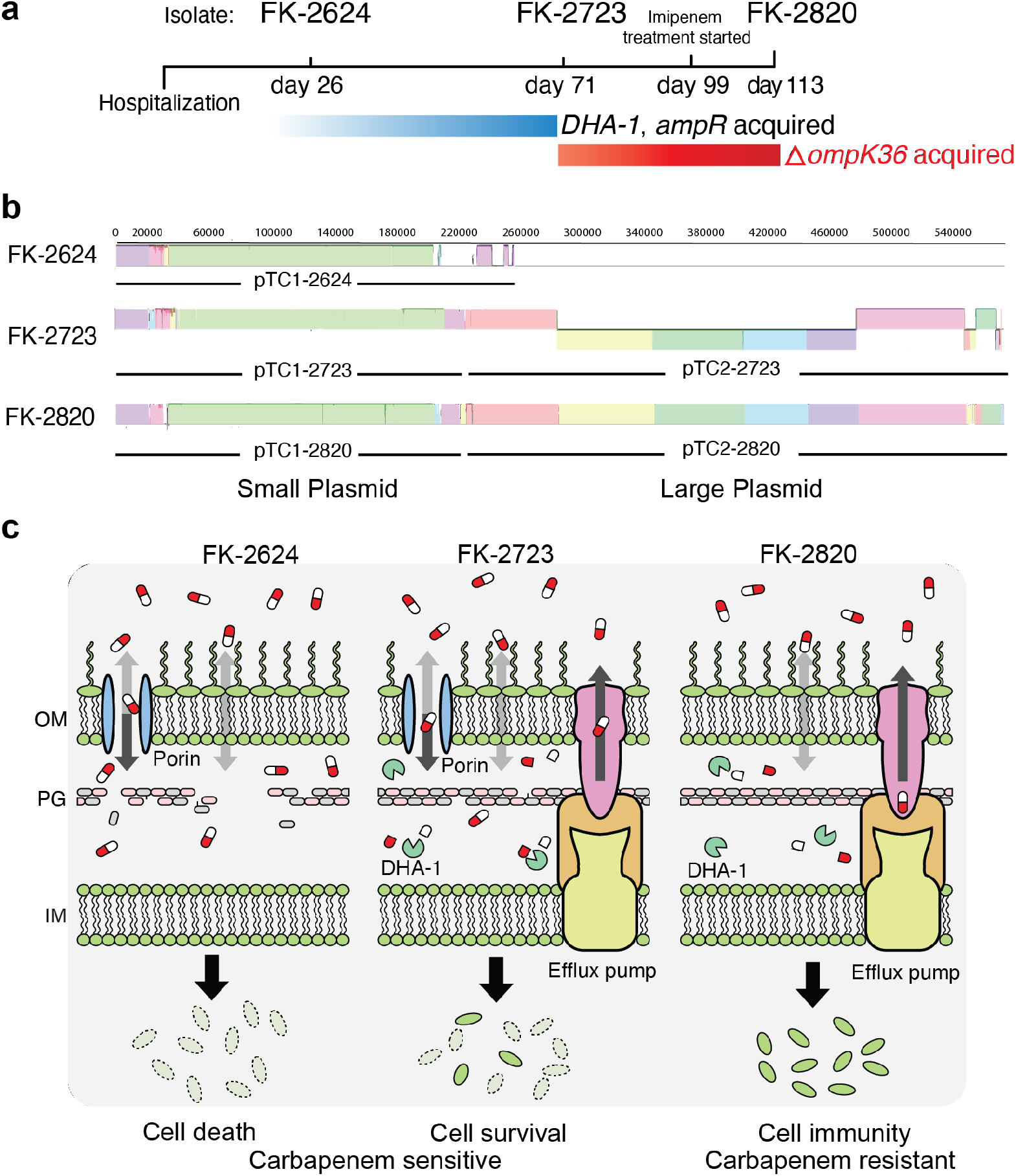
Mechanism of carbapenem resistance driven by a combination between membrane modifications and beta-lactamases. **a**, During the timeline of infection, beta-lactamases gene *bla*_DHA-1_and its regular *ampR* are acquired and the *ompK36* porin gene mutated. **b**, Sequence-based comparisons of the five plasmids identified in this study. Coloured blocks represent shared homologous regions that are free of genomic arrangements; their heights correspond to the average level of conservation in the NGS data. White coloured areas represent regions that are absent in comparison to other strains. Blocks above the centre line indicate forward orientation corresponding to the first sequence, while blocks below the line indicate reverse compliment orientation. We show detailed mapping of the small and large plasmids in Supplementary Fig. 1a. **c**, In susceptible strains (FK-2624, FK-2723), carbapenems enter the bacterial cell via porins in the outer membrane and inhibit the process of cell wall biogenesis in the periplasm. The acquisition of a plasmid-encoded beta-lactamase, and the acquisition of genes encoding a drug-efflux pump is not sufficient to deliver a carbapenem-resistant phenotype (FK-2723). However, with these genetic changes, a mutation in the porin gene to diminish the rate and extent of drug influx into the periplasm results in a carbapenem-resistant phenotype (FK-2820). An alignment of the porin gene *ompK36* is shown in Supplementary Fig. 1b.

The smaller plasmid shared by all three isolates showed ∼98% sequence identity across the three strains. Annotation of the plasmid-associated genes indicated that several resistance genes found in pTC1-2723 and pTC1-2820 were absent from pTC1-2624. Notably gene *bla*_*TEM-116*_ – encoding a broad-spectrum beta-lactamase – was acquired in the plasmid carried in FK-2723 (Fig. 2, **Supplementary Fig. 1a, Supplementary Table 2, 3)**.

The larger plasmids pTC2-2723 and pTC2-2820 showed ∼99% sequence identity to each other (Fig. 2b). The major difference between them is that pTC2-2820 was ∼4,000 nts longer due to duplication of four genes (*silC, silE, silR* and *silS*). These genes encode the subunits of an efflux pump that spans the outer and inner membrane, being composed of the SilA and SilB efflux RND transporter and the outer membrane protein SilC, with a functionally related periplasmic substrate-binding protein SilE (17) (Fig. 2c). Expression of the genes for these structural components of the pump are regulated by the sensory histidine kinase (SilS) and the ligand-sensing response regulator (SilR) (17). Increased gene copy for this efflux pump in pTC2-2820 could contribute to a change in the effective concentration of carbapenem in the periplasm of FK-2820 if the pump has any capacity to expel antibiotics (8, 9).

The plasmid acquired by FK-2723 and retained in FK-2820 also carries *bla*_DHA-1_, a gene encoding a beta-lactamase previously assessed as having no significant activity against carbapenems (18). The stepwise acquisition of efflux pumps and *bla*_DHA-1_ expression could have a combined effect to impact the drug concentration in the periplasm but is unlikely to produce a CRE phenotype. Rather, these genes predispose the strain through providing a “pre-AMR state” to the bacterial cells (Fig. 2c).

A compounding difference in FK-2820 relative to the two earlier strains comes from a 158 nt deletion in the chromosomal gene *ompK36*, which changes the reading frame and introduces a premature stop codon (Supplementary Fig. 1b). This explains the absence of detectable OmpK36 observed in immunoblots of FK-2820 (10). Mechanistically, the loss of porin function would decrease the entry of drug into the periplasm. Taken together, the WGS data depicts the coordination of an array of genes that potentiates a scenario for the evolution of a CRE phenotype within a single patient. We therefore sought to address how widespread this scenario might be in other patients and other strains of *Klebsiella*.

### Plasmid prevalence, diversity and carbapenem resistance mechanisms

To understand the prevalence and diversity of CRE mechanisms, we characterized 43 genomes representing 28 different multilocus sequence typing (MLST) types from *K. pneumoniae* strains and one *K. quasipneumoniae* strain. Details on sequencing, collection, and source of each of these strains is documented in Table S1. From these 43 strains, 42 of the genomes contained plasmid sequences which varied from ∼24,000 to ∼800,000 nt, with up to seven plasmids in one strain (Supplementary Table 3). Only *K. pneumoniae* strain RDAO01.1 appeared to be without a plasmid. While maximum likelihood phylogeny of their core genomes showed clustering by MLST types (Fig. 3), there was also significant evolutionary distance between the MLST types (Fig. 1b).

**Figure 3.**
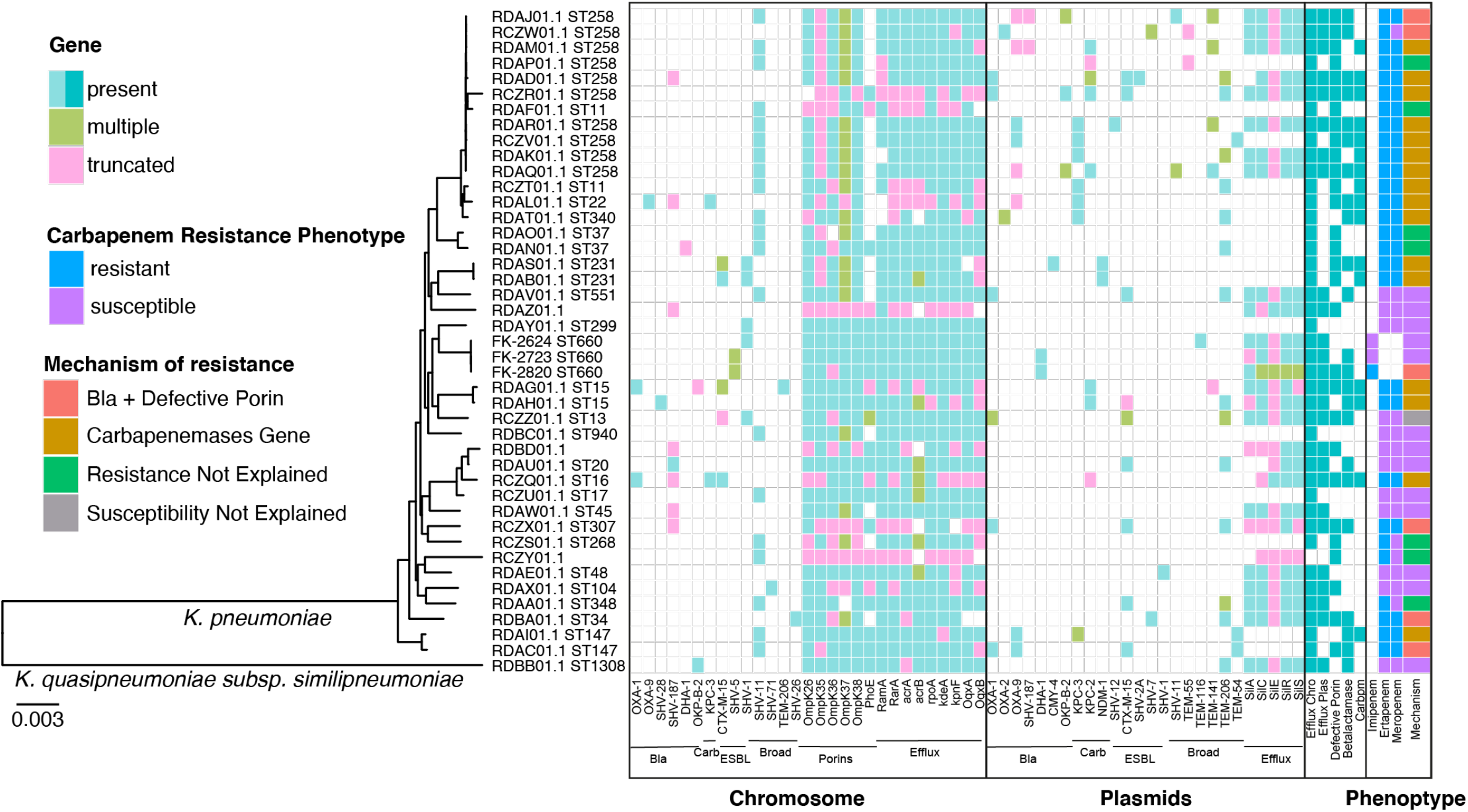
Resistance mechanisms of extensively characterised *Klebsiella* isolates. The maximum likelihood phylogeny was generated using the core genome of 164242 nt. The mechanism of resistance was deduced using presence or absence of genes. The labels in the bottom allow the identification of the gene groups as follows: Bla (beta-lactamases), Carb (carbapenemase), ESBL (extended spectrum beta-lactamases), Broad (broad spectrum beta-lactamases), Porins and Efflux genes. A distinguishing feature of FK-2820 is the truncation of OmpK36, with sequence analysis confirming that this consists of a deletion in the sequence that would cause synthesis of the polypeptide to be truncated by a premature stop codon. Scale bar represents nucleotide substitutions per site.

There is a current acceptance for a set of 289 genes that are known to contribute to AMR (6, 7, 9, 19), and these include the various isoforms of beta-lactamases, carbapenemases, porins and efflux pumps of interest in our study. We identified 45 of these AMR genes across the 43 genomes (Supplementary Table 4). Among the 14 carbapenem-susceptible strains, only RCZZ01 was found to have a defective porin gene (a gap of 720 nt in *ompK36*) and a plasmid-borne *bla*_OXA-1_ encoding a beta-lactamase, whereas the remainder had intact porin genes and no identifiable beta-lactamases (Fig. 3). Only 16 of the 43 *Klebsiella* genomes were found to encode a recognizable carbapenemase, either KPC-2, KPC-3, or NDM-1, and all these 16 isolates were documented as being of CRE phenotype.

The remaining 13 *Klebsiella* genomes came from strains documented as CRE, thus representing the non-CP CRE cohort of bacteria (Fig. 3). To understand the genes associated with the 13 strains that were non-CP CRE, we mapped the chromosomal and plasmid-borne resistance genes. Six of the non-CP CRE strains encoded plasmid-borne beta-lactamases and contained deletions in chromosomal genes encoding the porins OmpK26, OmpK35, OmpK36, OmpK37, OmpK38 or PhoE (Fig. 3, Supplementary Table 4). Consistent with patterns observed globally, the most detected defective porin was OmpK35 (7).

The other seven non-CP CRE strains did not express any recognizable beta-lactamase. While six of the seven strains showed defects in porin-encoding genes due to deletions, loss of porin function alone has not been associated with resistance to carbapenems. This comprehensive mapping of known resistance markers shows that almost as prevalent as the carbapenemase-producing CRE strains are non-CP CRE strains, where gene association between a defective porin and a plasmid-borne beta-lactamase gene is evident. The mapping also suggests that further gene associations contribute to the CRE phenotype.

### Gene association between plasmid-borne and chromosomal genes

To identify any new gene associations that may contribute CRE phenotypes we developed a computational workflow to analyse 69,512 predicted proteins across the 43 genomes (Fig. 4a and Supplementary Fig. 2). First, a t-distributed stochastic neighbour embedding (t-SNE) analysis was performed with a matrix of presence/absence/duplication of the predicted proteins (Fig. 4). The protein sequences common to all 43 genomes formed a “centroid” (Fig. 4b), whereas those protein sequences exclusive to specific MLST types clustered independently. To avoid evolutionary biases and to focus on acquired genes and phenotype related genotypes, we selected all putative proteins that deviated from both the centroid and the MLST-specific clusters. The resulting 3527 putative proteins thus represent the accessory genomes and uncommon variations.

**Figure 4.**
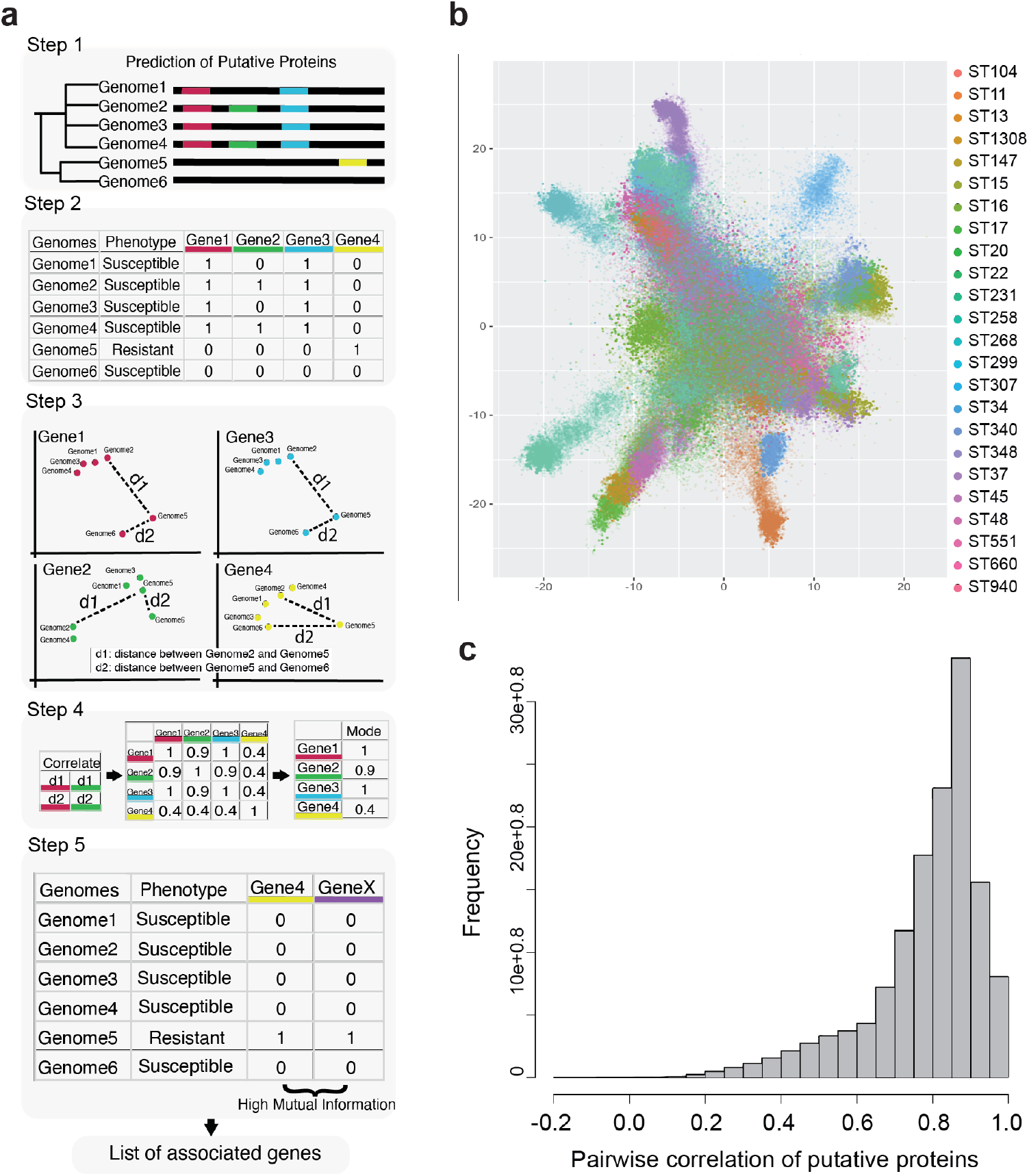
Identifying gene association in putative proteins. **a**, workflow to detect gene associations. Step 1, Putative proteins were annotated in the genomes, denotated as genes. Step 2, A matrix with the presence, absence and multiple copies of these genes was generated to represent the genotype of genome. Step 3, t-SNE was calculated per gene and the relative distance between genomes was represented in two dimensions. Step 4, The relative distances between genomes per gene was correlated pairwise through all gene combinations Step 5, the mode of the correlations per gene was calculated and genes with a mode less than 0.5 were selected for the next step. Step 5, Mutual information (MI) of all pairwise comparisons was calculated and genes with high MI (>0.9) were considered associated. A detailed illustration of the computational workflow is provided in Supplementary Fig. 2. **b**, Clustering of the putative proteins of 43 *K. pneumoniae* strains using tSNE. The colours represent the different MLST types. **c**, Histogram of the pairwise correlations of the distances between genomes in each putative protein. A high correlation can be observed for essential genes present in most strains, while putative proteins that represent the accessory genome and noncommon variations of genes will have lower correlations overall.

Pairwise Mutual Information (MI) was used to identify gene associations from the 3527 putative proteins selected. Any two proteins were considered highly associated when the presence or absence of one could predict the presence or absence of the other with a high MI score (>0.9). In six strains that produced beta-lactamases and contained defective porins, 65 genes were found to be associated with resistance (MI>0.9): 31 were exclusively in plasmids, 19 were carried in the chromosome, and 12 genes were not exclusively in either chromosome or plasmids, occurring seemingly at random either in plasmids or chromosomes (Table 1). Twenty-three of these 65 genes encode membrane proteins, showing enrichment of efflux pumps including the SilABC efflux pump and other resistance nodulation division (RND) transporters. In addition to the structural genes for efflux pumps, we identified the gene encoding the membrane protein PcoS, a sensor histidine kinase responsive to the intracellular concentration of copper, and regulating the expression of genes encoding metal ion pumps (20). The second most frequently associated genes were mobile elements. Thirteen genes identified were predicted to encode transposases and insertion genes. Two of these, IS*Kpn26* and IS*1*, are known to impact porin expression (6, 21).

**Table 1.**
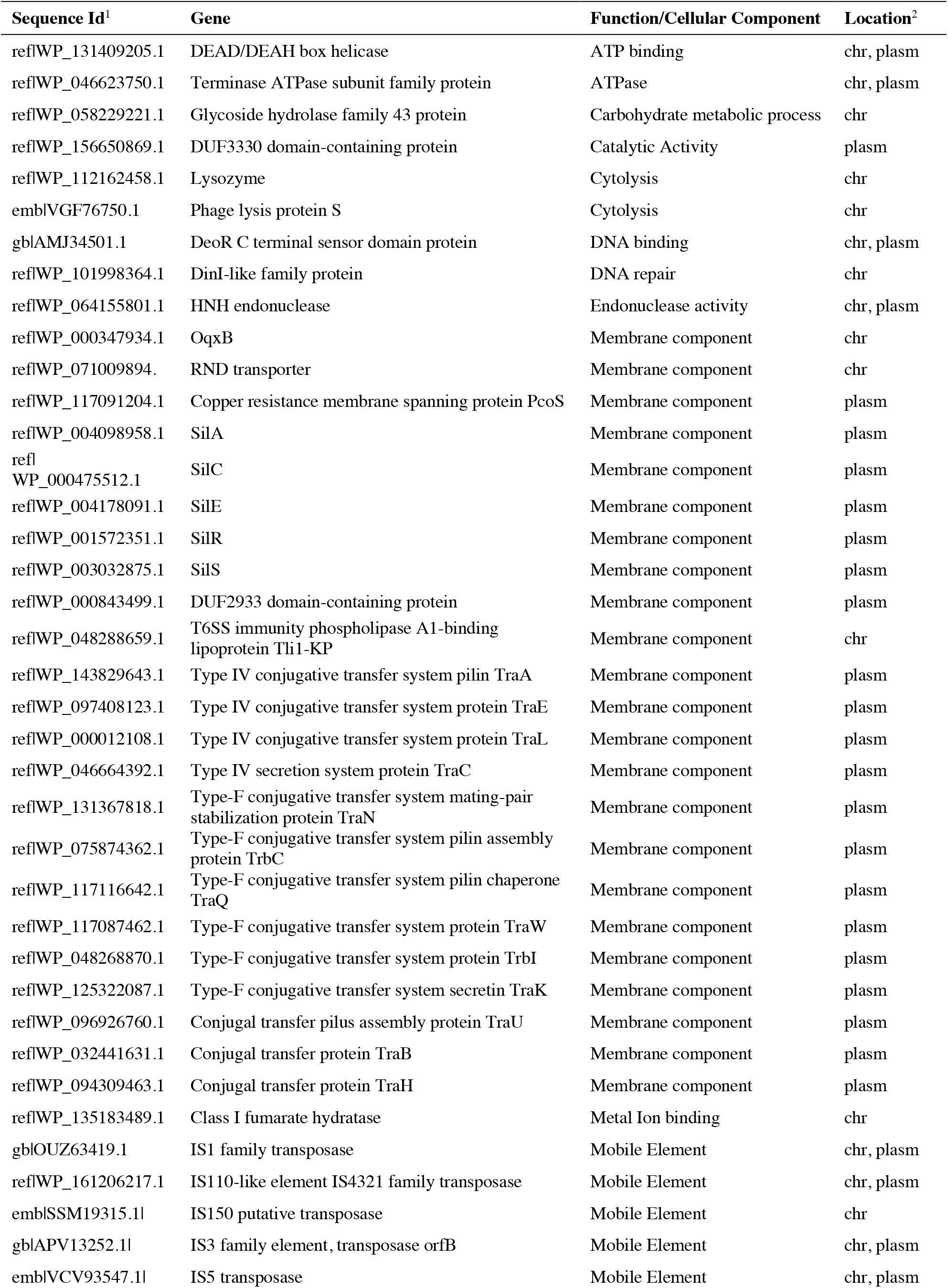
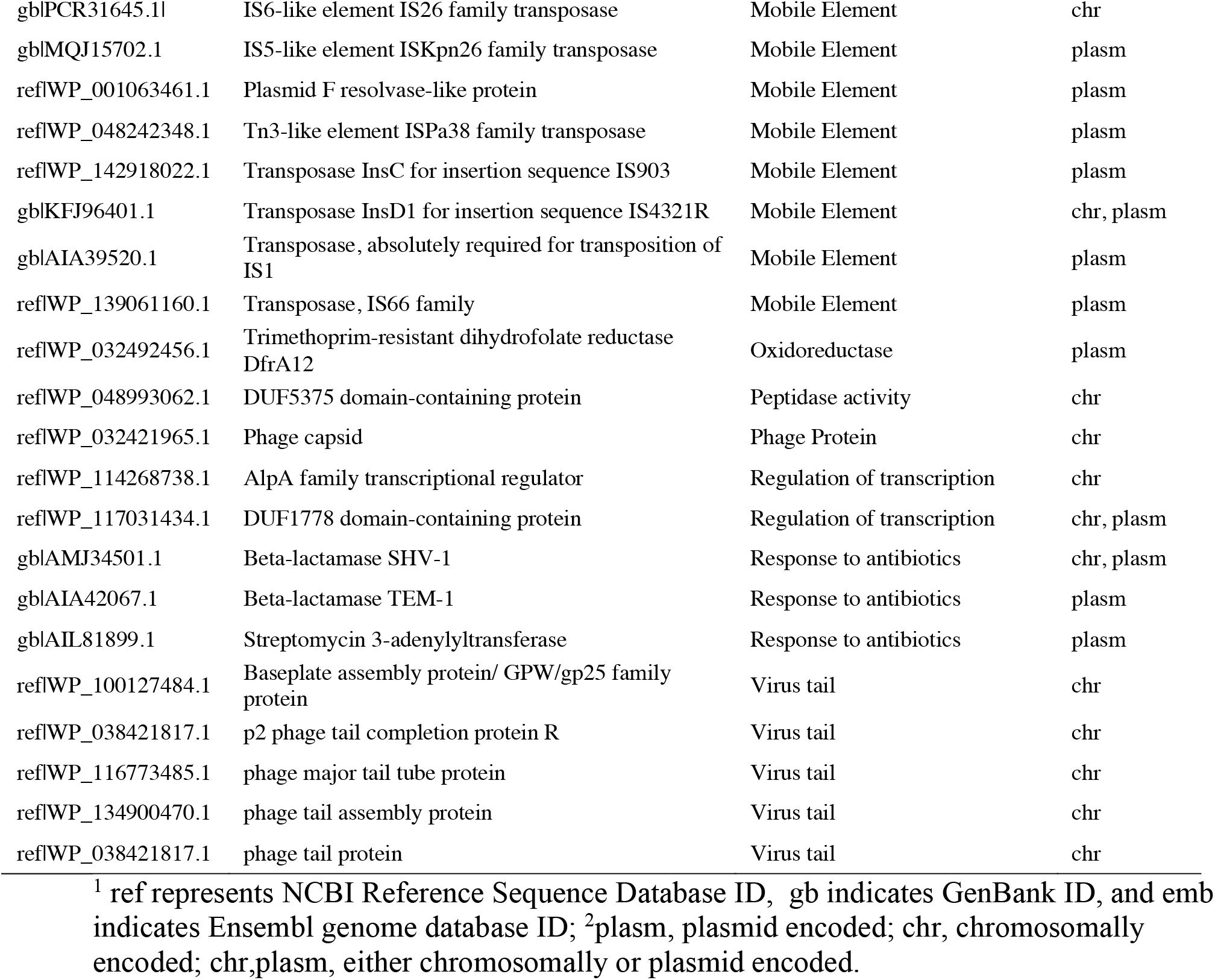
Genes found to be associated with non-CP CRE strains.

Focussing on the 7 CRE *Klebsiella* strains that had no known mechanism to explain their CRE phenotype was highly informative. We identified 224 genes that were not present in susceptible strains (Supplementary Table 5). These genes included efflux pumps (major facilitator superfamily (MFS) efflux pumps and RND efflux pumps), cell wall modifying enzymes (e.g. serine-type D-Ala-D-Ala carboxypeptidase) and detergent resistance mechanisms (e.g. benzoate/H(+) symporter BenE family transporter). High gene-association was found among 18 of these genes (14 chromosomal genes and 4 plasmid-borne genes) (Table 2). One example common to all 7 strains was association of the plasmid-borne gene *acrB*, and the porin-encoding chromosomal gene *lamB*. AcrB is a component of the AcrAB-TolC drug efflux pump, known to contribute to drug resistance (19). While the porin LamB is not generally recognized as contributing to carbapenem resistance in *Klebsiella* (7), it has been shown to contribute to CRE phenotypes in *Escherichia coli* (22).

**Table 2.**
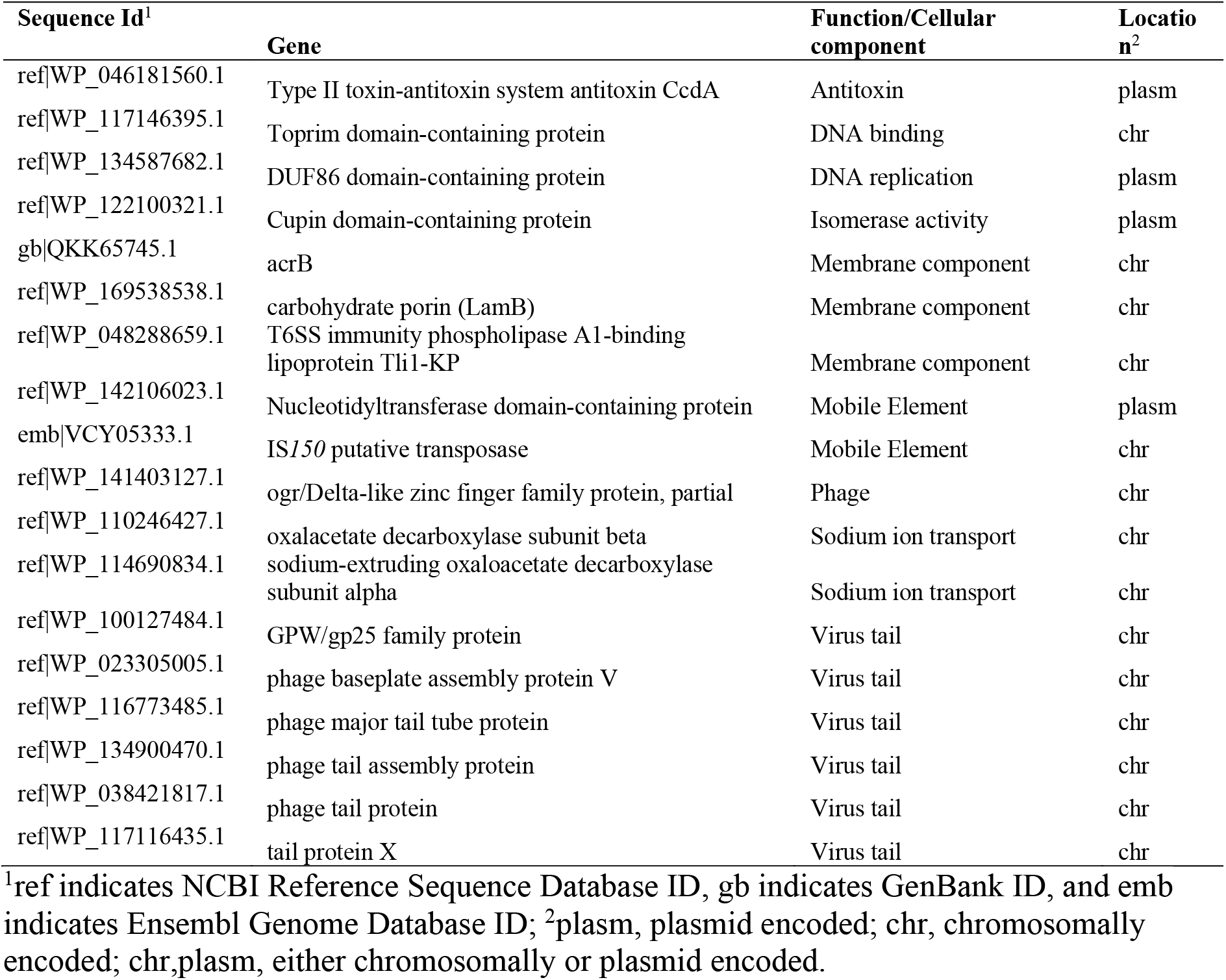
Genes found to be associated with strains that have unknown mechanisms of resistance to carbapenems.

In summary, we observed a synchronized presence of CRE phenotype-related genes in plasmids and on chromosomes in comparison to strains with carbapenem susceptible phenotypes. Our results suggest that for CRE strains the function of the gene carried by the plasmid was facilitated by the gene in the chromosome, and when these gene pairs are not present in concert, the strain will be susceptible to carbapenem treatment.

## DISCUSSION

While the evolution of CRE phenotypes in *Klebsiella* and other species of Enterobacteriaceae is on the rise, there remains many clinical cases where carbapenems can provide effective treatment against life-threatening infections. Rapid diagnosis to discriminate between CRE and carbapenem-sensitive phenotypes is in demand. WGS will be increasingly used as a basis for rapid diagnosis, but the gene signatures that determine a CRE phenotype are not as simple as once thought. Here, through systematic characterisation of complete *Klebsiella* genomes we identified three different groups of strains with multiple plasmid-mediated mechanisms of resistance to carbapenems: (i) the predictable strains with plasmid-borne genes encoding recognizable carbapenemases, (ii) strains with plasmids that encode other non-carbapenemase beta-lactamases in conjunction with chromosomal genes mutated to encode functionally inactivated porins, and (iii) other strains.

This ability of *Klebsiella* to use different mechanisms to mount a CRE phenotype adds to the complexity of its diagnosis. Only the first group of strains, those encoding recognizable carbapenemases, would be predicted as CRE by current genome-based diagnostics. Sixteen such strains were present in the 43 *Klebsiella* genomes analyzed. Three different carbapenemases were present (KPC-2, KPC-3 or NDM-1) and these genes were all present on plasmids. Carbapenems differ structurally from other beta-lactam drugs, accommodating an additional pyrrolidine ring in the active site that is not present in most other beta-lactams (23). Therefore, the carbapenemases KPC-2, KPC-3 or NDM-1 differ enough in structure from other beta-lactamases (24) that sequence feature-based diagnosis is robust, and the KPC-2 carbapenemase has become a dominant form diagnosed in many clinical investigations of CRE. Plasmids carrying *bla*_*KPC-2*_have contributed significantly to the success of *K. pneumoniae* ST258, which has been the dominant global clone of CRE *K. pneumoniae* (25).

While the conventional wisdom is that the beta-lactamase encoded by *bla*_DHA-1_ has no activity against carbapenems, such statements are based only on growth phenotypes and MIC evaluations. As an ESBL, the enzyme encoded by *bla*_DHA-1_ has broad substrate specificity for beta-lactams and has measurable activity against carbapenems *in vitro* (26). A recent review documents the diverse plasmids that have been shown to carry *bla*_DHA-1_ across bacterial species (27), and while carriage of *bla*_DHA-1_ alone does not confer CRE phenotypes, it can confer a reduced susceptibility to carbapenem treatment (26).

Porins are beta-barrel proteins in the outer membrane of Gram-negative bacteria and defects in genes encoding porins have been associated with carbapenem resistance (7, 18, 26). Several studies have assessed *Klebsiella* isolates phylogenetically and reported that *ompK35* is the most prevalent target of inactivating mutations in CRE *Klebsiella* (7). While few controlled studies have been done using isogenic strains expressing only one of the porins OmpK26, OmpK35, OmpK36, OmpK37, OmpK38 or PhoE, there is reason to expect that in a given strain of *Klebsiella*, there may be factors that dictate which of these proteins is most highly expressed and might therefore be most permissive to the influx of carbapenems into the periplasm. One such study concluded that all six porins can contribute somewhat to carbapenem transport into *Klebsiella*, but that OmpK35 appears to be the biggest contributor (7). Here we mapped potential inactivating mutations in OmpK26, OmpK35, OmpK36, OmpK37, OmpK38 and PhoE, and found that 29 strains from the 43 analysed had defective porins. Of these, 20 were defective in OmpK35, 13 defective in OmpK36, 5 defective in OmpK37 and 6 in OmpK38, with several strains having more than one porin truncated (Fig. 3). The mechanism by which porins are made defective range from SNV that incorporate premature stop-codons and those that inhibit antibiotic import whilst maintaining nutrient acquisition, to major changes mediated by insertion sequences such as IS*1* (21). IS*1* was found as an associated gene in the present study. Furthermore, 29 of the strains carried mutations in chromosomal genes encoding the porins OmpK26, OmpK35, OmpK36, OmpK37, OmpK38 or PhoE, with defects in *ompK35* being most prevalent and, of these, 22 also showed evidence of IS*1*.

In addition to the combined effect of defective porins and plasmid-encoded beta-lactamases, we found gene association data to suggest that expression of efflux pumps correlates to CRE phenotypes. Influx-efflux links such as those highlighted in Figure 2C would be one explanation for why efflux pump carriage is associated with CRE phenotypes (6). However, the control circuits governing efflux pump expression might also provide clues to understand the strains with no known resistance mechanism (28). The steady-state level of efflux pumps is controlled through drug-responsive transcription factors such as RamR and MarR. In their resting state, these repressors bind to the promoter regions of the transcriptional activators, *ramA* and *marA*, respectively. This prevents expression of the activators. In response to increased drug concentration, a conformational change is induced in RamR/MarR releasing them from their operator sites, and thereby derepressing *ramA* and *marA* to activate overexpression of *acrAB-tolC*. In a detailed case monitoring CRE evolution in *K. pneumoniae*, it was found that an SNV in RamR attenuated expression of *acrAB* genes and also *ompK35* (29). This study shows that second-site mutations can affect porin loss and provides a potential explanation to the third category of CRE phenotypes investigated here: those that did not encode a recognizable carbapenemase or a defective porin encoding gene.

t-SNE analysis has been used widely in single cell analysis; however, recently its potential to analyse diversity of populations and clustering of accessory genes has been explored, opening a diverse number of utilities for this method (30). Genome-wide analysis to identify gene associations and epistasis to elucidate interdependent genes that cause antibiotic resistance has proven to be challenging due to weak statistical power, the phylogenetic biases, and the need for large datasets (>1000 genomes) (31). Here we showed that clustering of pangenomes using t-SNE allowed the identification of gene associations that confer phenotypes of interest. We identified two categories of CRE *Klebsiella* that do not encode recognizable carbapenemases. In one of these categories, a combination of loss-of-function mutations in the strain’s major porin and plasmid carriage of an ESBL was detected. For this and the other non-carbapenemase CRE strains, an association of efflux pumps was detected. A comprehensive genome-based diagnostic for CRE will need to take into account all of these gene features.

A great leap forward in cancer diagnostics came with the concept that a given cancer evolves in a patient as a result of a series of stepwise mutations that result, phenotypically, in a set of precancerous states. This was based on early realizations (32, 33) and was thereafter shown to be a dominant and general mechanism for cancer progression (34). With advances in technology, single-cell studies have consolidated this view of pre-cancerous states evolving into recognizable cancers through multiple, step-wise, genetic changes that can be diagnosed early in the progression to cancer (35). Conceptually, the in-host evolution of carbapenem-resistance follows this same paradigm: a series of mutations results, phenotypically, with the bacterium manifesting a series of pre-AMR states. Neither acquisition of genes for beta-lactamase expression nor drug efflux pump expression alone makes the bacterium carbapenem-resistant. However, the ultimate mutation that makes a porin non-functional is the final stage in the progression to advanced drug-resistant infection. Diagnostics aimed at detailing these pre-AMR states could be advantageous in securing optimal treatment options, as is the case in cancer diagnostics (36).

## METHODS

### Genomic datasets

Three *Klebsiella* samples collected from a patient in the ICU of the First Affiliated Hospital in Wenzhou, China, during late 2015–early 2016 were sequenced using Illumina NovaSeq PE150 and deposited in GenBank with accession no. VIGL00000000, VIGM00000000 and VIGK00000000. In addition, we analysed the whole genomes of 40 *Klebsiella* genomes generated using nanopore sequencer (Oxford Nanopore Technologies) by John’s Hopkins Hospital Medical Microbiology Laboratory from clinical isolates collected during 2016 and 2017 (Project NCBI ID: PRJNA496461)(13).

### Genome Assembly

Sequence quality was analysed using FastQC (37). Primers were trimmed using FastX ToolKit (http://hannonlab.cshl.edu/fastx_toolkit/) and assembled using Unicycler v0.4 (38). *K. pneumoniae* strain ATCC 35657 (CP015134.1) was determined as the reference genome using MagicBlast v1.5 (39) and used for scaffolding in MeDuSa (40). The ordering of scaffolds was determined using MAUVE v02.2 (41). Kleborate v0.3.0 (42) was used to identify MLST types.

### Characterization of membrane components and resistance genes

The presence, absence, or truncation of porins, efflux and resistance genes in each genome were systematically characterized using the reference gene sequences as follows. First contigs belonging to plasmids and chromosomal genome were delineated using mlplasmids v1.0 (16). Genes prediction was performed using Prodigal v2.6.3 (43). The predicted genes were blasted to a database of genes of interest, including porin gene families, efflux, and resistance genes. Two hundred eighty-nine resistant, porin and efflux genes were tested. Forty-five genes were identified in the 43 genomes analysed (Supplementary Table 4). Gene alignments were generated in R, using packages “msa”, “reshape2”, “Biostrings” and “seqinr”, and parallelised using a perl script FromAssembly2gene.pl available in https://github.com/LPerlaza/Assembly2Gene. Additionally, plots illustrating gene organization and plasmid maps were generated using an in-house perl script GenePlot.pl.

### Phylogenetic analysis

Roary v3.11.2 (44) was used to align 597 complete *Klebsiella* genomes from the NCBI database (downloaded May 2020) and extract the core genomes. The core genome was used to generate a phylogenetical tree using RAxML v8.2.12 (45) with a general time reversible nucleotide substitution model with rate heterogeneity modelled with a gamma distribution (GTR+GAMMA). Branch supports were estimated using 1,000 bootstrap replicates.

### SNV analysis

Single nucleotide variants (SNVs) between each pairwise combination were detected using Parsnp (Harvest tool Suite, 1.1.2) (46). Gingr (Harvest tool Suite, 1.1.2) (46) was used to generate VCF files, and vcftools was used to summarize the SNVs counts. The expected number of SNVs between any two genomes was estimated assuming a mutation rate of 1 × 10^−7^ nucleotides per site per generation (47), thirty-six generations per day and the number of days between collections. SNVs in different isolates that were in the same positions, with the same nucleotide change were considered “shared mutations” (48).

### Comparative Genomics and gene association

To discover statistical association between predicted genes we developed a workflow as follows (Fig. 4a and Supplementary Fig. 2). First, we detected accessory genes and noncommon variations of genes (i.e. genes with deletions or insertions, duplicated genes) related with the resistant phenotype. We used a t-distributed stochastic neighbour embedding (t-SNE) analysis to identify clustering of genomes by putative proteins. All predicted pangenomic proteins with >90% coverage and >90% similarity were clustered using CD-HIT v4.8.1 (49), identifying 69,512 putative proteins in the 43 genomes analysed. Then a matrix of putative proteins presence/absence/multicopy was generated. T-SNE per protein was performed in R using package “Rtsne” v0.15 (https://github.com/jkrijthe/Rtsne) with the perplexity parameter of 10 determining the number of close neighbours in a group for 10 iterations and theta of 0.5.

Using the *X,Y coordinates* in the cartesian plane generated by the t-SNE analysis the distance between genomes per putative protein was calculated. Per each putative protein there is a set of distances between genomes. We considered the distances between genomes that have different resistant phenotypes (Resistant and Susceptible). The distance between genomes with the same phenotype were excluded. This avoided a biased correlation from being overpowered by distances between genomes with the same phenotypes, or capturing the signal of accessory genes related with other phenotypes. The correlation of different putative proteins pairwise distances reflected how similar those putative proteins group the genomes. Assuming that most putative proteins are not associating the gene by phenotypes but by phylogenetic closeness, the lowest correlated putative proteins were considered. The rationale behind this is that putative proteins that have similar pairwise distances (high correlation) group the genomes in clusters. Putative proteins that have lower correlations will deviate from the clustering that most putative proteins generate. This method was especially successful to identify different mechanisms of resistance because it does not seek perfect aggregations of genomes with different phenotypes but allows to detect putative proteins that deviate from the general clustering behaviour of the rest of putative proteins. The putative proteins with a mode lower than 0.5 were considered to deviate from the phylogenetically segregated clustering.

3527 putative proteins were found to be correlated with the centroid with less than 0.5. To determine gene association between plasmids and chromosome, these selected putative proteins were then correlated between each other using mutual information (MI), calculated using the R package “infotheo” v1.2.0. Four categories were used: susceptible strains (n=14), carbapenemases-producing strains (n=16), beta-lactamases producing strains with defective porins (n=6), and resistant strains with no known mechanism (n=7). Genes with high mutual correlation (>0.9) within these groups were considered associated genes that contribute to the phenotype. Additionally, these genes were investigated for functional annotation, gene ontology, and interaction. The functional annotations were determined by homology using Blast+ (50).

## Data Availability

The supplementary materials contain a list of sequence accession numbers. Code is available in https://github.com/LPerlaza/Assembly2Gene

## Acknowledgements

We thank Antonella Papa for critical discussions on the parallels in the evolution of cancer and infections. This work was supported by research grants from the Australian National Health and Medical Research Council (APP1092262), National Natural Science Foundation of China (no.81971986) and the Health Department of Zhejiang Province of the People’s Republic of China (no. 2011KYA106). These funding bodies provided funds for the purchase of consumption materials for the study but had no role in the design of the study and collection, analysis, and interpretation of data and writing of the manuscript.

## Author Contributions

T.L. and V.D. conceived the study. L.P.-J., T.Z., T.L., and V.D. designed the experiments. L.P.-J. performed the bioinformatics, and data analysis. T.C. and Y.Z. performed clinical isolation, resistance characterization, and genome sequencing. L.P.-J.

C.J.S. and J.J.W. performed characterization and validation experiments and analyzed carbapenem resistance mechanisms in case study. L.P.-J., T.L., and V.D. wrote the manuscript.

## Competing Interest Statement

The authors declare no competing interests.

## Supplementary materials

**Supplementary Fig. 1. a**, Comprehensive mapping of the small and large plasmids found in *K. pneumoniae* within-host isolates. **b**, Alignment of *ompK36* genes from the Wenzhou within-host isolates showing a premature stop codon in FK-2820.

**Supplementary Fig. 2**. Illustration of the workflow to detect associations. Flowchart of all the *in-silico* processes to detect gene associations. First, the genome contigs were classified as chromosomal or plasmid sequences and the putative proteins were predicted. The putative proteins were clustered by >90% coverage and >90% similarity. These clusters then were used to generate a matrix with presence, absence, and multiple copies of the putative proteins. Putative proteins were visualized using tSNE and the distance between genomes were calculated. Those distances were then correlated between putative proteins. If the mode of correlations per each putative protein was less than 0.5 the putative protein was selected. The selected putative proteins were grouped by phenotype and were then correlated using Mutual Information (MI). Putative proteins with more than 0.9 of MI were considered correlated.

**Supplementary Table 1**. Summary of genomes analysed

**Supplementary Table 2**. *In silico* detection of resistance

**Supplementary Table 3**. Annotated genes for all plasmids.

**Supplementary Table 4**. Genotype of all resistance genes, porins, and efflux pump genes analysed, including descriptive information of the genes studied for each genome, including classification of presence, truncated, absence, as well as SNPs, insertions, deletions and duplications.

**Supplementary Table 5**. Genes present only in strains with unknown mechanisms of resistance.

## Notes

### Competing Interest Statement

The authors have declared no competing interest.

### Author Declarations

All of the investigation protocols in this study were approved by the Ethics Committee of the First Affiliated Hospital of Wenzhou Medical University (ethical number 2019-75). Informed consent was waived because this retrospective study focused on the bacterial isolates and had no impact on interventions to the patient.

## References

1. W. H. O. WHO (2017) Global priority list of antibiotic-resistant bacteria to guide research, discovery, and development of new antibiotics.

2. Y. Zhang et al., Evolution of hypervirulence in carbapenem-resistant Klebsiella pneumoniae in China: a multicentre, molecular epidemiological analysis. J Antimicrob Chemother 75, 327–336 (2020).

3. P. Nordmann, T. Naas, L. Poirel, Global spread of Carbapenemase-producing Enterobacteriaceae. Emerg Infect Dis 17, 1791–1798 (2011).

4. D. van Duin et al., Molecular and clinical epidemiology of carbapenem-resistant Enterobacterales in the USA (CRACKLE-2): a prospective cohort study. Lancet Infect Dis 10.1016/S1473-3099(19)30755-8 (2020).

5. S. M. Drawz, R. A. Bonomo, Three decades of beta-lactamase inhibitors. Clin Microbiol Rev 23, 160–201 (2010).

6. M. H. Nicolas-Chanoine, N. Mayer, K. Guyot, E. Dumont, J. M. Pages, Interplay Between Membrane Permeability and Enzymatic Barrier Leads to Antibiotic-Dependent Resistance in Klebsiella Pneumoniae. Front Microbiol 9, 1422 (2018).

7. A. Rocker et al., Global Trends in Proteome Remodeling of the Outer Membrane Modulate Antimicrobial Permeability in Klebsiella pneumoniae. mBio 11 (2020).

8. V. B. Srinivasan, B. B. Singh, N. Priyadarshi, N. K. Chauhan, G. Rajamohan, Role of novel multidrug efflux pump involved in drug resistance in Klebsiella pneumoniae. PLoS One 9, e96288 (2014).

9. V. B. Srinivasan, G. Rajamohan, KpnEF, a new member of the Klebsiella pneumoniae cell envelope stress response regulon, is an SMR-type efflux pump involved in broad-spectrum antimicrobial resistance. Antimicrob Agents Chemother 57, 4449–4462 (2013).

10. X. Tian et al., First description of antimicrobial resistance in carbapenem-susceptible Klebsiella pneumoniae after imipenem treatment, driven by outer membrane remodeling. BMC Microbiol 20, 218 (2020).

11. R. F. Silva et al., Pervasive sign epistasis between conjugative plasmids and drug-resistance chromosomal mutations. PLoS Genet 7, e1002181 (2011).

12. M. Lukacisinova, B. Fernando, T. Bollenbach, Highly parallel lab evolution reveals that epistasis can curb the evolution of antibiotic resistance. Nat Commun 11, 3105 (2020).

13. P. D. Tamma et al., Applying Rapid Whole-Genome Sequencing To Predict Phenotypic Antimicrobial Susceptibility Testing Results among Carbapenem-Resistant Klebsiella pneumoniae Clinical Isolates. Antimicrob Agents Chemother 63 (2019).

14. A. Drouin et al., Predictive computational phenotyping and biomarker discovery using reference-free genome comparisons. BMC Genomics 17, 754 (2016).

15. M. Nguyen et al., Developing an in silico minimum inhibitory concentration panel test for Klebsiella pneumoniae. Sci Rep 8, 421 (2018).

16. S. Arredondo-Alonso et al., mlplasmids: a user-friendly tool to predict plasmid- and chromosome-derived sequences for single species. Microb Genom 4 (2018).

17. S. Silver, Bacterial silver resistance: molecular biology and uses and misuses of silver compounds. FEMS Microbiology Reviews 27, 341–353 (2003).

18. Z. Kis, A. Toth, L. Janvari, I. Damjanova, Countrywide dissemination of a DHA-1-type plasmid-mediated AmpC beta-lactamase-producing Klebsiella pneumoniae ST11 international high-risk clone in Hungary, 2009-2013. J Med Microbiol 65, 1020–1027 (2016).

19. V. B. Srinivasan, A. Mondal, M. Venkataramaiah, N. K. Chauhan, G. Rajamohan, Role of oxyRKP, a novel LysR-family transcriptional regulator, in antimicrobial resistance and virulence in Klebsiella pneumoniae. Microbiology (Reading) 159, 1301–1314 (2013).

20. G. P. Munson, D. L. Lam, F. W. Outten, T. V. O’Halloran, Identification of a copper-responsive two-component system on the chromosome of Escherichia coli K-12. Journal of Bacteriology 182, 5864–5871 (2000).

21. J. Vandecraen, M. Chandler, A. Aertsen, R. Van Houdt, The impact of insertion sequences on bacterial genome plasticity and adaptability. Crit Rev Microbiol 43, 709–730 (2017).

22. U. Choi, C. R. Lee, Distinct Roles of Outer Membrane Porins in Antibiotic Resistance and Membrane Integrity in Escherichia coli. Frontiers in Microbiology 10, 1–9 (2019).

23. T. Naas et al., Beta-lactamase database (BLDB) - structure and function. J Enzyme Inhib Med Chem 32, 917–919 (2017).

24. K. Bush, G. A. Jacoby, Updated functional classification of beta-lactamases. Antimicrob Agents Chemother 54, 969–976 (2010).

25. C. R. Lee et al., Global Dissemination of Carbapenemase-Producing Klebsiella pneumoniae: Epidemiology, Genetic Context, Treatment Options, and Detection Methods. Front Microbiol 7, 895 (2016).

26. H. Mammeri, H. Guillon, F. Eb, P. Nordmann, Phenotypic and biochemical comparison of the carbapenem-hydrolyzing activities of five plasmid-borne AmpC beta-lactamases. Antimicrob Agents Chemother 54, 4556–4560 (2010).

27. C. Hennequin, V. Ravet, F. Robin, Plasmids carrying DHA-1 beta-lactamases. Eur J Clin Microbiol Infect Dis 37, 1197–1209 (2018).

28. N. Maurya, M. Jangra, R. Tambat, H. Nandanwar, Alliance of Efflux Pumps with beta-Lactamases in Multidrug-Resistant Klebsiella pneumoniae Isolates. Microb Drug Resist 25, 1155–1163 (2019).

29. M. Aihara et al., Within-host evolution of a Klebsiella pneumoniae clone: selected mutations associated with the alteration of outer membrane protein expression conferred multidrug resistance. J Antimicrob Chemother 76, 362–369 (2021).

30. K. Abudahab et al., PANINI: Pangenome Neighbour Identification for Bacterial Populations. Microb Genom 5 (2019).

31. B. Schubert, R. Maddamsetti, J. Nyman, M. R. Farhat, D. S. Marks, Genome-wide discovery of epistatic loci affecting antibiotic resistance in Neisseria gonorrhoeae using evolutionary couplings. Nat Microbiol 4, 328–338 (2019).

32. H. M. Temin, Evolution of cancer genes as a mutation-driven process. Cancer Res 48, 1697–1701 (1988).

33. E. R. Fearon, B. Vogelstein, A genetic model for colorectal tumorigenesis. Cell 61, 759–767 (1990).

34. C. Tomasetti, B. Vogelstein, G. Parmigiani, Half or more of the somatic mutations in cancers of self-renewing tissues originate prior to tumor initiation. Proc Natl Acad Sci U S A 110, 1999–2004 (2013).

35. N. McGranahan, C. Swanton, Clonal Heterogeneity and Tumor Evolution: Past, Present, and the Future. Cell 168, 613–628 (2017).

36. J. Beane, J. D. Campbell, J. Lel, J. Vick, A. Spira, Genomic approaches to accelerate cancer interception. Lancet Oncol 18, e494–e502 (2017).

37. S. Andrews, F. Krueger, A. Seconds-Pichon, F. Biggins, S. Wingett (2015) FastQC. A quality control tool for high throughput sequence data. Babraham Bioinformatics.

38. R. R. Wick, L. M. Judd, C. L. Gorrie, K. E. Holt, Unicycler: Resolving bacterial genome assemblies from short and long sequencing reads. PLoS Comput Biol 13, e1005595 (2017).

39. G. M. Boratyn, J. Thierry-Mieg, D. Thierry-Mieg, B. Busby, T. L. Madden, Magic-BLAST, an accurate RNA-seq aligner for long and short reads. BMC Bioinformatics 20, 405 (2019).

40. E. Bosi et al., MeDuSa: a multi-draft based scaffolder. Bioinformatics 31, 2443–2451 (2015).

41. A. C. Darling, B. Mau, F. R. Blattner, N. T. Perna, Mauve: multiple alignment of conserved genomic sequence with rearrangements. Genome Res 14, 1394–1403 (2004).

42. K. L. Wyres et al., Identification of Klebsiella capsule synthesis loci from whole genome data. Microb Genom 2, e000102 (2016).

43. D. Hyatt et al., Prodigal: prokaryotic gene recognition and translation initiation site identification. BMC Bioinformatics 11, 119 (2010).

44. A. J. Page et al., Roary: rapid large-scale prokaryote pan genome analysis. Bioinformatics 31, 3691–3693 (2015).

45. A. Stamatakis, RAxML version 8: a tool for phylogenetic analysis and post-analysis of large phylogenies. Bioinformatics 30, 1312–1313 (2014).

46. T. J. Treangen, B. D. Ondov, S. Koren, A. M. Phillippy, The Harvest suite for rapid core-genome alignment and visualization of thousands of intraspecific microbial genomes. Genome Biol 15, 524 (2014).

47. R. Sanjuan, M. R. Nebot, N. Chirico, L. M. Mansky, R. Belshaw, Viral mutation rates. J Virol 84, 9733–9748 (2010).

48. C. J. Worby, M. Lipsitch, W. P. Hanage, Shared Genomic Variants: Identification of Transmission Routes Using Pathogen Deep-Sequence Data. Am J Epidemiol 186, 1209–1216 (2017).

49. L. Fu, B. Niu, Z. Zhu, S. Wu, W. Li, CD-HIT: accelerated for clustering the next-generation sequencing data. Bioinformatics 28, 3150–3152 (2012).

50. C. Camacho et al., BLAST+: architecture and applications. BMC Bioinformatics 10, 421 (2009).

