## Supplementary Figure 1 for "Stepwise evolution of carbapenem-resistance, captured in patient samples and evident in global genomics of *Klebsiella pneumoniae*"

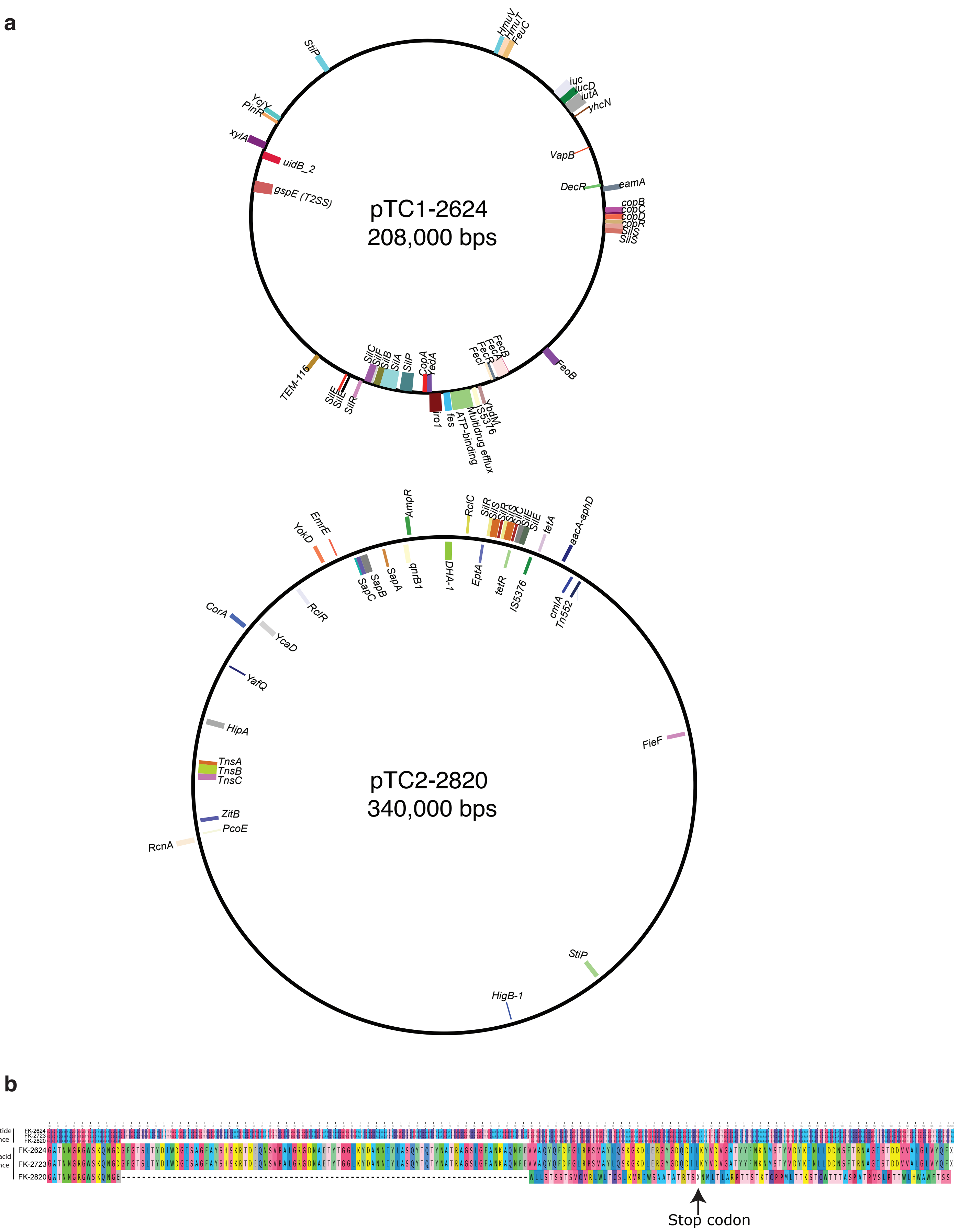


**Supplementary Fig. 1. a**, Comprehensive mapping of the small and large plasmids found in K. pneumoniae within-host isolates. **b**, Alignment of *OmpK36* genes from the Wenzhou within-host isolates showing a premature stop codon in FK-2820 resulting in frameshift an truncation.
