## Supplementary Figure 2 for "Stepwise evolution of carbapenem-resistance, captured in patient samples and evident in global genomics of *Klebsiella pneumoniae*"

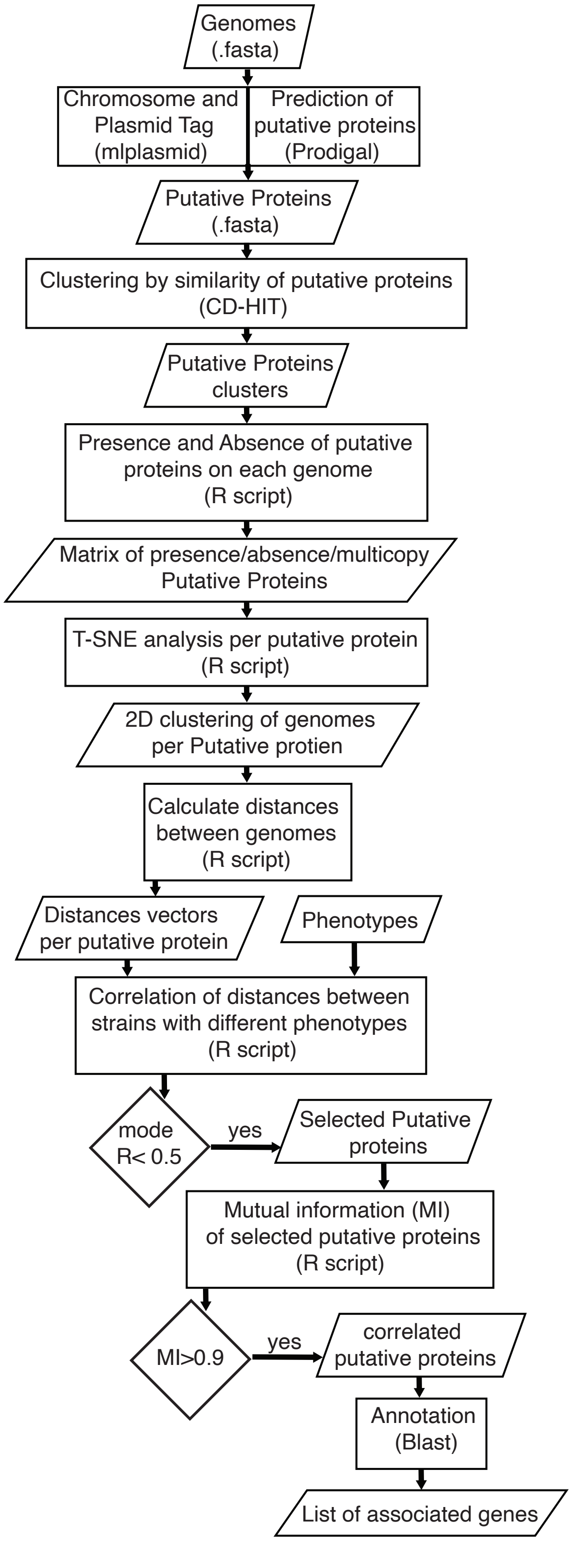


**Supplementary Fig. 2. Illustration of the workflow to detect associations.** Flowchart of all the in-silico processes to detect gene associations. First, the genome contigs were classified as chromosomal or plasmid sequences and the putative proteins were predicted. The putative proteins were clustered by >90% coverage and >90% similarity. These clusters then were used to generate a matrix with presence, absence, and multiple copies of the putative proteins. Putative proteins were visualized using tSNE and the distance between genomes were calculated. Those distances were then correlated between putative proteins. If the mode of correlations per each putative protein was less than 0.5 the putative protein was selected. The selected putative proteins were grouped by phenotype and were then correlated using Mutual Information (MI). Putative proteins with more than 0.9 of MI were considered correlated.
